# Genome-wide association study of prostate-specific antigen levels in 392,522 men identifies new loci and improves cross-ancestry prediction

**DOI:** 10.1101/2023.10.27.23297676

**Authors:** Thomas J Hoffmann, Rebecca E Graff, Ravi K Madduri, Alex A Rodriguez, Clinton L Cario, Karen Feng, Yu Jiang, Anqi Wang, Robert J Klein, Brandon L Pierce, Scott Eggener, Lin Tong, William Blot, Jirong Long, Louisa B Goss, Burcu F Darst, Timothy Rebbeck, Joseph Lachance, Caroline Andrews, Akindele O Adebiyi, Ben Adusei, Oseremen I Aisuodionoe-Shadrach, Pedro W Fernandez, Mohamed Jalloh, Rohini Janivara, Wenlong C Chen, James E Mensah, Ilir Agalliu, Sonja I Berndt, John P Shelley, Kerry Schaffer, Mitchell J Machiela, Neal D Freedman, Wen-Yi Huang, Shengchao A Li, Phyllis J Goodman, Cathee Till, Ian Thompson, Hans Lilja, Dilrini K Ranatunga, Joseph Presti, Stephen K Van Den Eeden, Stephen J Chanock, Jonathan D Mosley, David V Conti, Christopher A Haiman, Amy C Justice, Linda Kachuri, John S Witte

## Abstract

We conducted a multi-ancestry genome-wide association study of prostate-specific antigen (PSA) levels in 296,754 men (211,342 European ancestry; 58,236 African ancestry; 23,546 Hispanic/Latino; 3,630 Asian ancestry; 96.5% of participants were from the Million Veteran Program). We identified 318 independent genome-wide significant (p≤5e-8) variants, 184 of which were novel. Most demonstrated evidence of replication in an independent cohort (n=95,768). Meta-analyzing discovery and replication (n=392,522) identified 447 variants, of which a further 111 were novel. Out-of-sample variance in PSA explained by our genome-wide polygenic risk scores ranged from 11.6%-16.6% in European ancestry, 5.5%-9.5% in African ancestry, 13.5%-18.2% in Hispanic/Latino, and 8.6%-15.3% in Asian ancestry, and decreased with increasing age. Mid-life genetically-adjusted PSA levels were more strongly associated with overall and aggressive prostate cancer than unadjusted PSA. Our study highlights how including proportionally more participants from underrepresented populations improves genetic prediction of PSA levels, offering potential to personalize prostate cancer screening.

## Introduction

Prostate-specific antigen (PSA) is a protein encoded by the *KLK3* gene and secreted by the prostate gland^1–3^. PSA levels are often elevated in those with prostate cancer (PCa); however, elevated levels can also be caused by other factors, such as benign prostatic hyperplasia (BPH), local inflammation or infection, prostate volume, age, and germline genetics^4–8^. PSA screening for PCa was approved by the Food and Drug Administration in 1994, but it is unclear if the benefits for PCa-specific mortality reduction outweigh the harms from overdiagnosis and treatment of clinically insignificant disease^9–12^. Previous work estimates 20-60% of screen-detected PCas are overdiagnoses (i.e., PCa that would not otherwise clinically manifest, or result in PCa-related death^13^), other work has suggested that 229 individuals would need to be invited to screen and 9 would need to be diagnosed to prevent one death^14^, and the United States^15^, Canada^16^, and the United Kingdom^17^ recommend against universal population-based screening. If PSA levels could be adjusted for an individual’s predisposition in the absence of PCa, the specificity (to reduce overdiagnosis) and sensitivity (to prevent more deaths) of screening could be improved.

Twin studies estimate PSA heritability at 40-45%^18,19^, and genome-wide heritability estimates at 25%-30%^20^ suggesting that incorporating genetic factors may improve screening. Recent work from our group based on 85,824 European ancestry and 9,944 non-European ancestry men found that genetically-adjusted PSA (i.e., the PSA measure inflated or deflated due to an individual’s genetic variants) most improved the discrimination of PSA screening for aggressive tumors^20^. In that work, we identified 128 genome-wide significant variants that explained up to 7% of PSA variation in European ancestry, suggesting that many more PSA loci remain.

Genome-wide polygenic risk scores (PRSs) explained up to 10% in European ancestry; however, the PRSs were less predictive in other groups, especially African ancestry (1-3%). Additional variant discovery with larger, more diverse cohorts could provide novel insights into the genetic architecture of PSA and further improve PCa screening.

## Results

### Composition of discovery and replication cohorts

Our discovery population consisted of 296,754 men without PCa from 9 cohorts: 211,342 European ancestry (71.2%), 58,236 African ancestry (19.6%), 23,546 Hispanic/Latino (7.9%), and 3,630 Asian ancestry (1.2%). None had been included in previous PSA GWASs. We present genotype platform details in **Table S1**, demographics in **Table S2**, and quality control (QC) metrics in **Table S3**. The pooled mean age at PSA measurement across the discovery cohorts was 57.4 (standard deviation [SD]=9.6), and the pooled median PSA was 0.84. The Million Veteran Program (MVP) comprised 96.5% of the discovery cohort. For replication, we utilized results from 95,768 independent individuals from previous work^20^, including 85,824 European ancestry, 3,509 African ancestry, 3,098 Hispanic/Latino, and 3,337 Asian ancestry individuals (**Table S3**). **Figure 1** summarizes our analytical workflow and describes cohort ancestry compositions.

**Figure 1.**
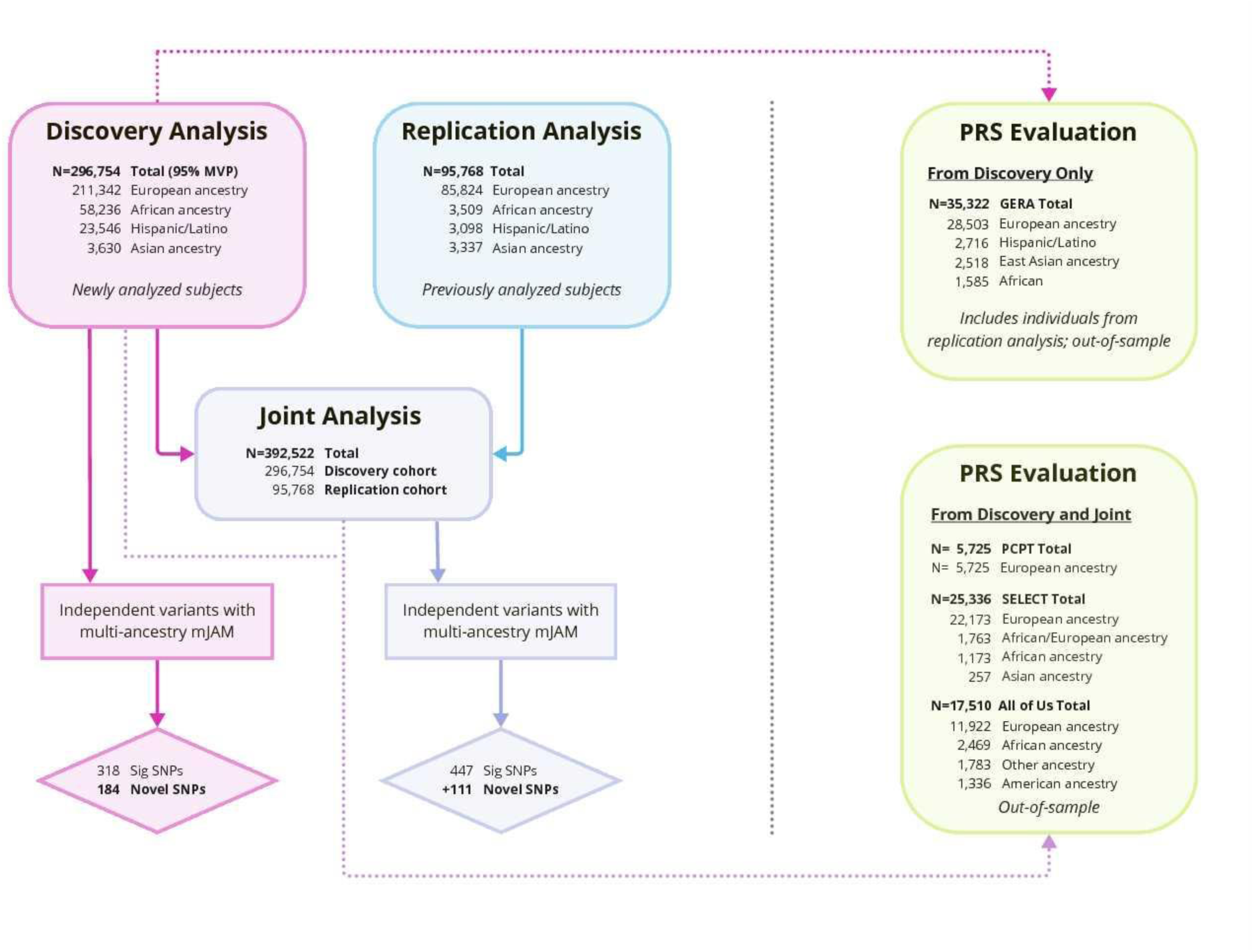
Flowchart describing the Precision PSA project analysis workflow and ancestry compositions of the discovery, replication, and joint meta-analysis cohorts. The discovery GWAS analysis revealed 318 genome-wide significant (p<5e-8) SNPs associated with PSA levels, of which 184 were novel. The joint analysis (consisting of the discovery and replication cohorts) revealed 447 genome-wide significant SNPs associated with PSA levels, of which an additional 111 were novel. Both discovery and joint GWAS results were used to develop PRSs for PSA, which were then evaluated in GERA (when out-of-sample), PCPT, and SELECT.

### Discovery GWAS analysis of PSA-associated variants

In our discovery cohorts, we identified 318 independent genome-wide significant variants (264 European, 51 African, 17 Hispanic/Latino, and 2 Asian) in a multi-ancestry analysis of log-transformed PSA levels (**Figure 2**, **Figure S1**, **Table S4**, **Table S5**) that used multiple reference panels to account for different ancestries (see **Methods**). Among them, 184 independent variants selected by mJAM^21^ were novel (see **Methods**). Of the novel variants, 57 replicated at a Bonferroni level (p<0.05/184=0.00027, same direction of effect on PSA), an additional 80 replicated at p<0.05 (and the same direction), 43 demonstrated the same effect direction (but p>0.05), and four showed no indication of replication (effect in the opposite direction). On average, compared to the non-replicated variants, the replicated variants had slightly larger effect sizes (mean β=0.30 versus 0.27) and were slightly more precise (mean standard error=0.0039 versus 0.0042).

**Figure 2.**
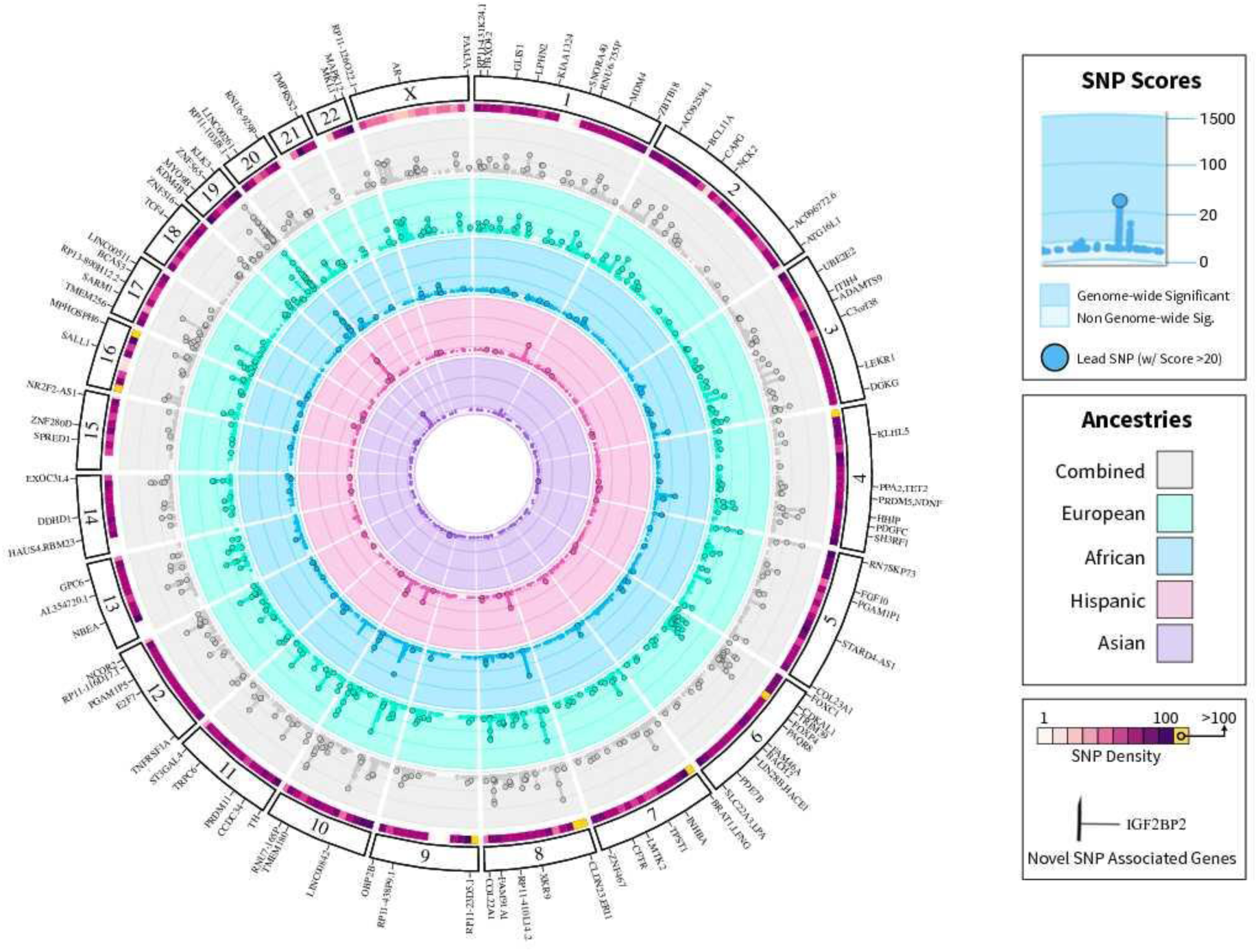
Circos plot showing PSA GWAS significant variants by chromosome from the discovery cohort. Concentric tracks are colored based on results from individual ancestries, with gray indicating results from the overall discovery meta-analysis. The top 100,000 GWAS SNPs (with the smallest p-values) per ancestry are shown as points; larger circled points indicate the 318 genome-wide significant variants (p<5e-8; 184 of which were novel) from the overall discovery analysis in all ancestries. SNP density in 10Mbp bins from the overall analysis is shown as a heatmap above the overall track. The outermost ring displays genes associated with novel discovery PSA SNPs.

Of the 184 variants that were novel in the multi-ancestry analysis, 112 were genome-wide significant in the European ancestry discovery cohort, eight were genome-wide significant in the African ancestry cohort, and none were genome-wide significant in Asian or Hispanics/Latino individuals (likely due to low sample size; **Figure S2**). Of the eight in African ancestry individuals, only two variants were frequent enough (see **Methods**) to be assessed in other ancestry groups: rs2071041 (*ITIH4*; β_African_=0.0237, 95% CI=0.0152-0.0322, p_African_=4.9e-8) that was also genome-wide significant in European ancestry individuals (β_European_=0.0180, 95% CI=0.0124 to 0.0235, p_European_=2.6e-10, minor allele frequency (MAF)=23.7%), and rs1203888 (*LINC00261*; β_African_=-0.0423, 95% CI=-0.0539 to -0.0307, p_African_=8.7e-13) that was not significant in European ancestry individuals (p>.05, MAF=0.8%). The latter variant showed similar magnitude of effect but was not Bonferroni significant in discovery Hispanic/Latino individuals (β_Hispanic/Latino_=-0.0748, 95% CI=-0.120 to -0.0297, p_Hispanic/Latino_=0.0012, MAF=3.1%) and was not significant in discovery Asian ancestry (p>.05, MAF=3.5%) or the replication cohorts (p>.05) (**Table S4**). The remaining six African ancestry variants were too rare to be assessed in European ancestry individuals. The variant rs184476359 (*AR*, multi-ancestry discovery β=-0.0590, 95% CI=-0.0774 to -0.0406, p=3.4e-10; replication β=-0.0870, 95% CI=- 0.1370 to -0.0371, p=6.3e-4) was common in African ancestry individuals (MAF=17.7%), less common in Hispanic/Latinos (MAF=1.1%), and not adequately polymorphic to be imputed in Asian individuals. Three variants in genes that encode PSA (rs76151346, β_African_=0.0821, 95% CI=0.0577 to 0.107, p_African_=4.6e-11, *KLK3*; rs145428838, β_African_=0.224, 95% CI=0.165 to 0.284, p_African_=1.4e-13, *KLK3*; rs182464120 β_African_=-0.213, 95% CI=0.278 to 0.147, p_African_=2.0e-10, *KLK2*) exclusively imputed in African ancestry individuals (all MAF<5%, two <1%) did not exhibit strong evidence of replication in African ancestry individuals(p>0.05). The remaining two variants identified in African ancestry (rs7125654, β_African_=-0.384, 95% CI=-0.0489 to -0.0279, p_African_=7.0e-13; rs4542679, β_African_=0.0422, 95% CI=0.0288 to 0.0557, p_African_=7.9e-10), were more common (MAF>5%) but also did not replicate (p>0.05). Further, rs7125654 (*TRPC6*) was less common in Latinos, but more common in Asian ancestry, and rs4542679 (*RP11-345M22.3*) was less common in Latinos and not adequately polymorphic in Asians.

We next tested for effect size differences across ancestry groups for the 184 novel variants. Only one variant, rs12700027 (*BRAT1*/*LFNG*, I^2^=84.8, p=0.00019), demonstrated heterogeneity that was Bonferroni-significant (p<0.05/184=0.00027). The variant had a strong discovery effect in European ancestry individuals (β=0.0327, 95% CI=0.0247 to 0.0407, p=1.2e-15, MAF=0.10), but was not significant in other groups (African ancestry β=0.0131, 95% CI=-0.0190 to 0.0452, p=0.42, MAF=0.021; Asian β=-0.176, 95% CI=-0.0203 to 0.0797, p=0.027, MAF=0.021; Hispanic/Latino β=-0.0102, 95% CI=-0.0326 to 0.0121, p=0.37, MAF=0.120). In our replication cohort, the variant nominally (i.e., p<0.05) replicated (p=0.0065, β=0.0175, 95% CI=0.00491 to 0.0302; European ancestry p=0.003, β=0.0327, 95% CI=0.0247 to 0.0407) and showed no statistically significant evidence of differences across ancestry groups (I^2^=0.0, p=0.44), although sample sizes for detecting differences were smaller.

In-silico assessment of potential functional features revealed that 20 of the novel variants (10.8%) were prostate tissue expression quantitative trait loci (eQTLs), and another 65 (35.3%) were eQTLs in other tissues (**Table S4**). Five novel variants were missense and predicted to be deleterious, with Combined Annotation Dependent Depletion (CADD) scores >20 (**Table S4**): rs11556924 in *ZC3HC1*, which regulates cell division onset; rs74920406 in *ELAPOR1*, a transmembrane protein; rs2229774 in *RARG*, a gene in the hormone receptor family; rs113993960 (delta508) in *CFTR*, a causal mutation for cystic fibrosis^22^; and rs2991716 upstream of LOC101927871. An additional 11 variants were predicted to have high pathogenicity (CADD scores >15;**Table S4**).

### Replication analysis of previously-reported variants in the discovery cohort

When we tested 128 previously identified variants^20^ in our discovery cohort, 106 (82.8%) replicated at a genome-wide significance level, an additional 15 replicated (11.7%) at a Bonferroni level (p<0.05/128=0.00039), an additional 6 replicated at p<0.05 (4.7%), and one variant flipped effect direction (**Table S6**). Replication was highest for European ancestry, likely due to sample size, with 94 variants (73%) reaching genome-wide significance, an additional 22 variants (17.2%) meeting a Bonferroni-corrected level, and 8 (6.3%) additional variants meeting p<0.05 (**Table S6**). Replication rates within African ancestry, our next largest group, were lower: 16 (12.5%) were genome-wide significant, 26 others (20.3%) met a Bonferroni level, an additional 39 (30.5%) had p<0.05, 32 additional (25.0%) were in the same direction, and 15 (11.7%) were in the opposite direction. Estimated rates were similar for Hispanic/Latino and lowest for Asian populations. Lastly, 16 of the 128 known variants showed heterogeneity across the four groups (Bonferonni corrected p<0.05/128=0.00039).

### Joint meta-analysis of discovery and replication cohorts

In the multi-ancestry analysis including the discovery and replication cohorts, we identified 447 independent variants (409 European, 56 African, 22 Hispanic/Latino, 6 Asian, including 46 in >1 group; **Figure 3, S1**; **Table S7**, **S8**). Among the 111 variants that were novel even relative to discovery alone, none showed evidence of ancestry effect size differences (p>0.05/111=0.00045). Fifty-six (50.4%) of the 111 were genome-wide significant in European ancestry individuals, but none were genome-wide significant in a non-European ancestry group (**Table S8**). Allele frequencies and effect sizes of the novel variants largely followed those expected by power curves (**Figure 4**).

**Figure 3.**
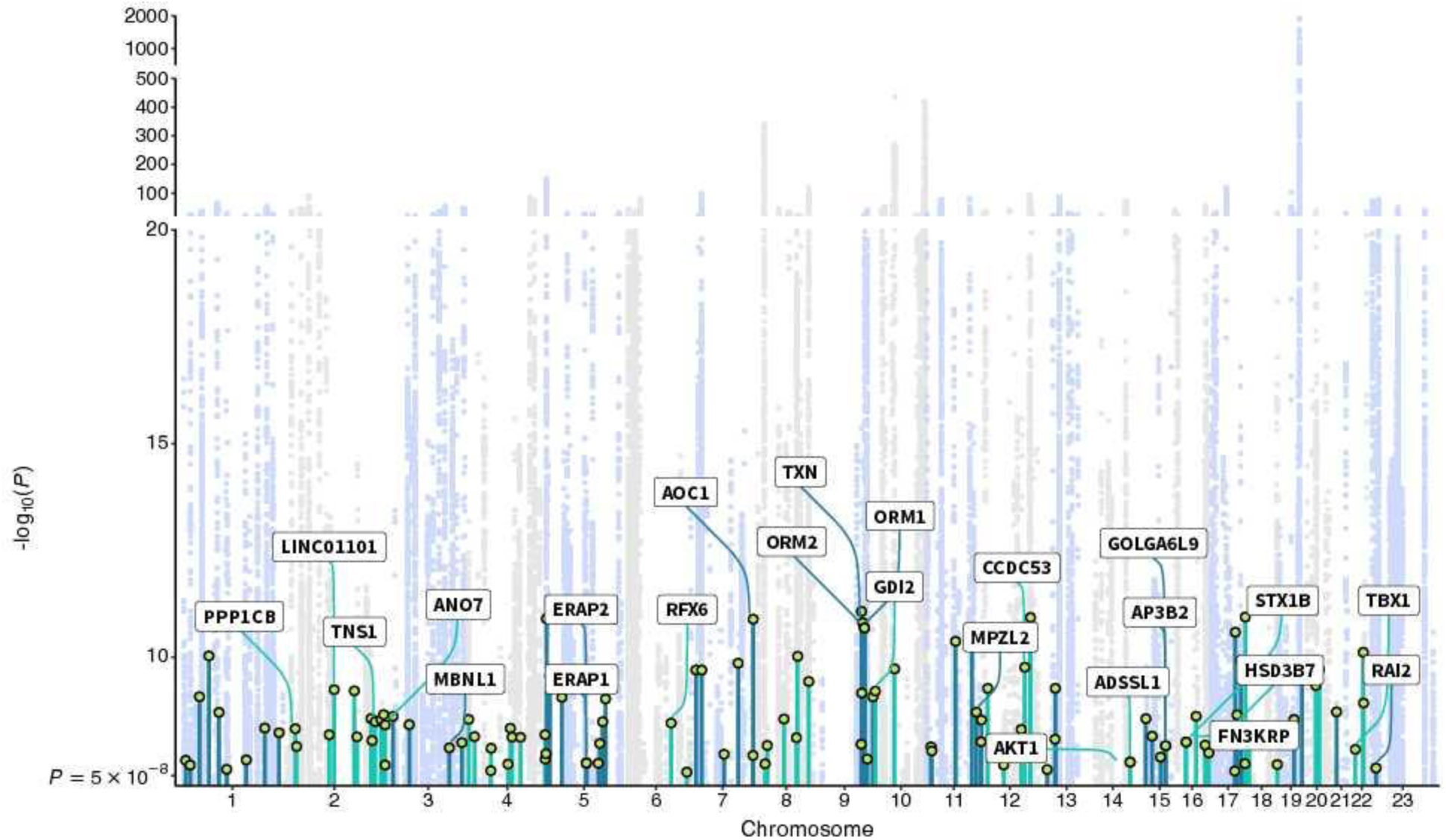
Manhattan plot showing results from the joint multi-ancestry meta-analysis of the discovery (n=296,754) and replication (n=95,768) studies. Only genome-wide significant associations (p<5e-8) are plotted. The joint analysis detected 447 independent genome-wide significant PSA-associated SNPs. These included 111 novel variants that were conditionally independent from previous findings and the discovery only analyses in each study alone (indicated by the circles). Gene labels are given for variants with CADD>15 and/or variants that are prostate tissue eQTLs.

**Figure 4.**
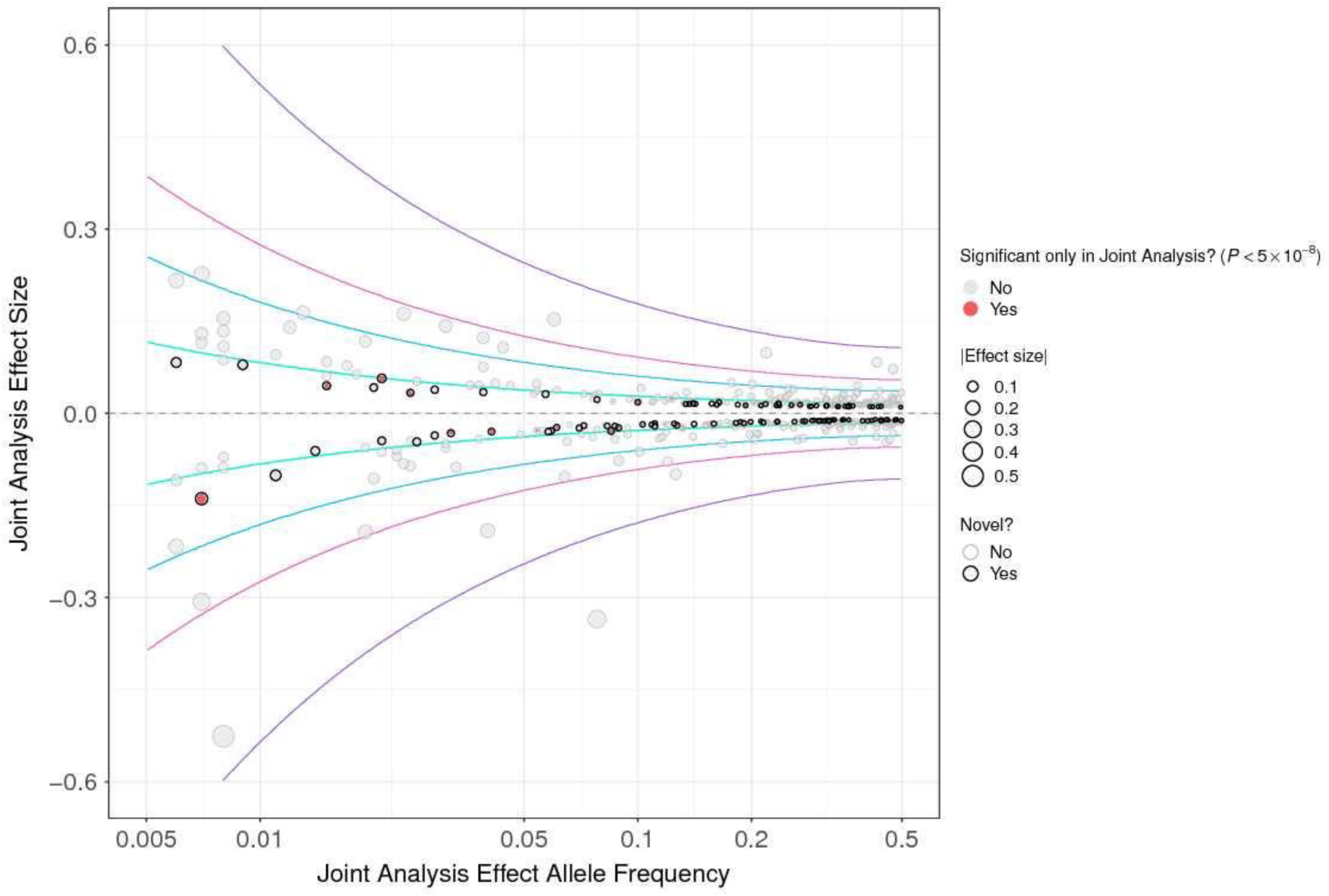
Plot of the relationship between minor allele frequency and estimated effect sizes for PSA GWAS significant variants. Each point represents one of the 447 independent genome-wide significant SNPs identified in our mJAM multi-ancestry GWAS joint meta-analysis. The SNP effect sizes are expressed in ln(PSA) per minor allele. The curves indicate the hypothetical detectable SNP effect sizes for a given minor allele frequency, assuming statistical power of 80%, α=5e-8 (genome-wide significant), and the sample size of each of our populations here (297,166 European ancestry, N=61,745 African ancestry, 6,967 Asian ancestry, 26,644 Hispanic).

In the joint meta-analysis, 12 (10.8%) novel variants were prostate tissue eQTLs, and 50 (45.0%) additional were eQTLs for other tissues. Two were missense substitutions (**Table S7**): rs1049742 in *AOC1* and rs74543584 in *MPZL2*. Three additional novel variants had CADD scores>15: rs1978060, an eQTL for *TBX1* in prostate tissue; rs339331 an eQTL for *FAM162B* in adipose tissue; and rs57580158, an intergenic variant with evidence of conservation.

### Medication sensitivity analysis

The UK Biobank (UKB) had medication information available to conduct a sensitivity analysis to see whether excluding individuals taking medications that could affect PSA levels (i.e., 5-alpha reductase inhibitors and testosterone) would impact our results. For PSA-associated variants, our primary results in the UK Biobank (UKB) were highly correlated (R=0.93, **Figure S3**) with the sensitivity analyses, suggesting these medications did not impact our results.

### Out-of-sample PSA variance explained by PRSs

We evaluated different strategies for constructing PRSs for PSA levels first using results from our discovery cohort (see **Methods**). For testing these PRSs, four cohorts of men without PCa were out-of-sample: Kaiser Permanente’s Genetic Epidemiology Research on Adult Health and Aging (GERA) cohort, the Selenium and Vitamin E Cancer Prevention Trial (SELECT),^23^ the Prostate Cancer Prevention Trial (PCPT),^24^ and All of Us (AOU)^25^.

In GERA, PRS_318_, constructed from the 318 conditionally-independent genome-wide significant variants in the multi-ancestry meta-analysis, generally had higher variance explained when using longitudinal measurements, rather than earliest PSA value, with 13.9% (95% CI=13.1%- 14.6%) in European ancestry (n=35,322), 13.1% (95% CI=10.6%-15.6%) in Hispanics/Latinos (n=2,716), 9.3% (95% CI=6.8%-12.0%) in African ancestry (n=1,585), and 9.0% (95% CI=7.0%-11.4%) in East Asian ancestry (n=2,518). The variance explained in the other three cohorts was ∼3-6% lower depending on the group (**Table S9**).

Expanding to a genome-wide approach, PRS-CSx (PRS_CSx-disc_; included more than genome-wide significant variants (1,070,230 SNPs; see **Methods**) and resulted in improved predictive performance. The variance explained increased to 16.6% (95% CI=15.9%-17.5%) in men of European ancestry and 18.2% (95% CI=15.4%-20.8%) in Hispanic/Latino men (**Figure 5A**, **Table S9**). The relative increase was largest in East Asian ancestry, with variance explained reaching 15.3% (95% CI=12.7%-18.1%), and smallest in African ancestry, with variance explained 8.5% (95% CI=6.1%-11.0%).

**Figure 5.**
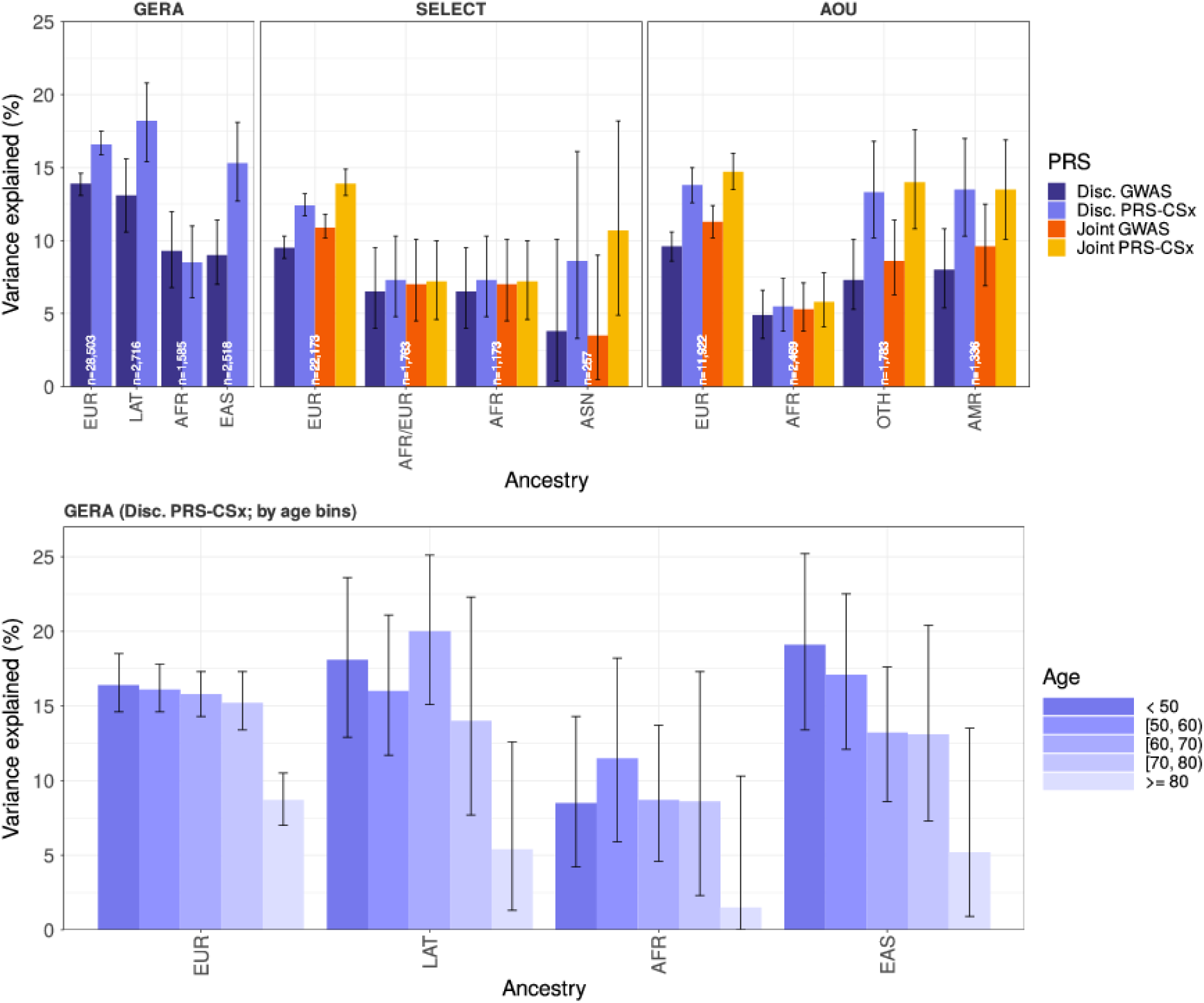
Variance in PSA levels explained by PRSs. We constructed PRSs for PSA from our discovery cohorts to allow assessment in GERA and our PRS validation cohorts (PCPT and SELECT). We also constructed PRSs from the joint meta-analysis of discovery and replication cohorts, with assessment in our validation cohorts. The PRSs were based on the multi-ancestry identified conditionally independent genome-wide significant variants using mJAM and on a multi-ancestry genome-wide score using PRS-CSx. The genome-wide score generally performed better than the genome-wide significant score. The variance explained by genome-wide PRSs **(A)** was up to 16.9% in Europeans, 18.6% in Hispanics/Latinos, 9.5% in Africans, and 15.3% in East Asians, and **(B)** decreased as age increased.

Second, we developed PRSs for PSA using the results from the joint GWAS meta-analysis (n=392,522), which combined the discovery meta-analysis with previously published results from Kachuri et al^20^. These scores were validated in PCPT, SELECT, and AOU, but not GERA, which was included in the previously published meta-analysis and was therefore not out-of-sample.

For the independent genome-wide significant PRSs, PRS_318_ explained 9.5% (8.8%-10.3%) of variation in baseline PSA levels in SELECT European ancestry (n=22,173), while PRS_447_ (from the 447 conditionally independent genome-wide significant variants identified in the joint meta-analysis) explained 10.9% (10.2%-11.8%) of the variance, which exceeded the 8.5% (95% CI=7.8%-9.2%) variance explained by PRS_128_ (from the 128 independent variants described in our prior GWAS of 95,768 men^20^). Variance explained in PCPT European ancestry (n=5,725) was slightly lower. In AOU European ancestry (n=11,922), variance explained was slightly higher, with PRS_128_ explaining 8.6% (95% CI=7.7%-9.6%), PRS_318_ explaining 9.6% (95% CI=8.6%-10.6%), and PRS_447_ explaining 11.3% (95% CI 10.2%-12.4%). We did not observe an appreciable change in differences across cohorts after removing individuals with BPH, a condition known to influence PSA levels. However, variance explained was slightly higher in all populations (<0.5% higher), albeit with overlapping CIs (**Table S9**).

Among SELECT African ancestry (n=1,173), PRS_128_ explained 3.4% (95% CI=1.6%-5.8%), PRS_318_ explained 6.5% (95% CI=4.0%-9.5%), and PRS_447_ explained 7.0% (95% CI=4.5%-10.1%); PRS_447_ more than doubled variance explained by PRS_128_. AOU African ancestry (n=2,471) estimates were 1-2% smaller.

A genome-wide PRS-CSx (PRS_CSx-joint_) compared to PRS_CSx-disc_ modestly increased variance explained by roughly 1-1.5% in European ancestry in PCPT (11.6%, 95% CI=10.0%-13.1%), SELECT (13.9%, 95% CI=13.1%-14.9%), and AOU (14.7%, 95% CI=13.5%-16.0%). PRS_CSx-joint_ also improved ∼3% upon the previously published PRS from the Kachuri et al.^20^ paper estimated here to be 8.6% in PCPT and 10.4% in SELECT for PRS-CSx (PRS_CSx-Kachuri_, **Table S9**). Among men of African ancestry in SELECT, PRS_CSx-joint_ showed no improvement (7.2%, 95% CI=4.6%-10.0%) over PRS_CSx-disc_, while variance explained in AOU increased by 0.3% (5.8%, 95% CI=4.1%-7.8%). Notably, PRS_CSx-joint_ yielded a substantial improvement upon the previously published PRS_CSx-Kachuri_ estimates^20^ of 1.64% in SELECT, although still under half of that in European ancestry.

In SELECT European ancestry individuals, PRS-CSx explained 13.9% of variation, while PRS_447_ explained 10.9% of the variation. Assuming that variation explained is nested between these approaches, we estimate 78.4% (=10.9%/13.9%) of PRS-CSx variation may be explained by PRS_447_. This is expected since information across the different PRSs overlaps, and the initial genome-wide significant SNPs from our large-scale GWAS are the most informative for explaining variation in PSA levels.

Third, we examined how the variance in PSA levels explained by the PRSs varied by age. These analyses were performed in GERA to have a large enough sample size in each age group and used PRS_CSx-disc_ to provide out-of-sample estimates. The estimated variance explained by the PRSs decreased with increasing age in all GERA ancestry groups, albeit with somewhat wide confidence intervals (**Figure 5B, Table S10**). For example, PRS_CSx-disc_ explained 16.4% (95% CI 14.6%-18.5%) of variation in PSA levels among European ancestry individuals <50y, and this decreased to 8.7% (95% CI 7.0%-10.5%) for men ≥80y.

Finally, for PRSs constructed from genome-wide significant independent variants, the variance explained when using weights corresponding to effect sizes from the multi-ancestry meta-analysis was almost always equal to or higher than variance explained when using ancestry-specific weights (**Table S10**). This was observed both for the discovery (PRS_318_) and joint meta-analysis (PRS_447_). The few instances where the variance explained was estimated lower almost always had <1% difference and wide confidence intervals around the estimate (i.e., the smallest sample sizes likely had unstable estimates).

### Relationship of PSA PRSs with PCa aggressiveness using Gleason score

In GERA, we performed a case-only analysis to examine the association between PSA PRS_CSx,disc_ (the out-of-sample PRSs with the highest variance explained) and Gleason score. Results were consistent with previous work that suggested that screening bias decreases the likelihood of identifying high-grade disease, whereby men with higher PRS values (indicating a genetic predisposition to higher constitutive PSA levels) are more likely to be biopsied, but less likely to have high grade disease^20^; we found that in European ancestry cases, an SD increase in PRS_CSx-disc_ was inversely associated with Gleason 7 (OR=0.78, 95% CI=0.73-0.84, p=1.2e- 13) and ≥8 (OR=0.71, 95% CI=0.64-0.79, p=6.2e-10) compared to Gleason ≤6. Other ancestry groups had similar estimated ORs, though they were not always statistically significant, likely owing to sample size (**Table S11**; e.g., African ancestry Gleason 7 (OR=0.88, 95% CI=0.67- 1.17, p=0.39) and ≥8 (OR=0.65, 95% CI=0.43-0.99, p=0.043)).

### Impact of genetically-adjusted PSA on prostate biopsy eligibility

We examined how PRS_CSx,disc_ would have changed biopsy recommendations for cases and controls, according to age-specific thresholds in GERA (see **Methods**). In European ancestry individuals who had negative biopsies (i.e., controls, n=2378), 16.0% with unadjusted PSA levels that exceeded age-specific thresholds for biopsy were reclassified to ineligible for biopsy. Among controls with PSA levels that did not indicate biopsy, 2.4% were reclassified to biopsy eligible, resulting in a control net reclassification improvement (NRI) of 13.6% (95% CI=12.2%- 15.0%; **Figure 6A**, **Table S12**). In individuals with positive biopsies (i.e., cases; n=2,358), 3.9% were reclassified to eligible, while 13.1% were reclassified to ineligible, resulting in a case NRI of -9.2% (95% CI=-10.3% to -8.0%). Of cases who became ineligible, 71.1% had Gleason scores ≤7, as compared to 56.5% who remained eligible (although we note that some of these men may have had biopsies for reasons other than their PSA level, such as abnormal digital rectal exam or strong family history). In African ancestry controls (n=110), 16.0% were reclassified to ineligible, while 2.4% were reclassified to eligible, resulting in an NRI of 3.6% (95% CI=0.1% to 7.1%; **Figure 6B**). In African ancestry cases (n=310), 5.2% were reclassified to eligible and 6.8% were reclassified to ineligible, resulting in an NRI of -1.6% (95% CI=3.0% to -0.2%). Other groups are shown in **Figure S4** with details in **Table S12**. We additionally obtained eight years of additional follow up on the 78 controls in all groups now classified as eligible; three were later diagnosed with PCa.

**Figure 6.**
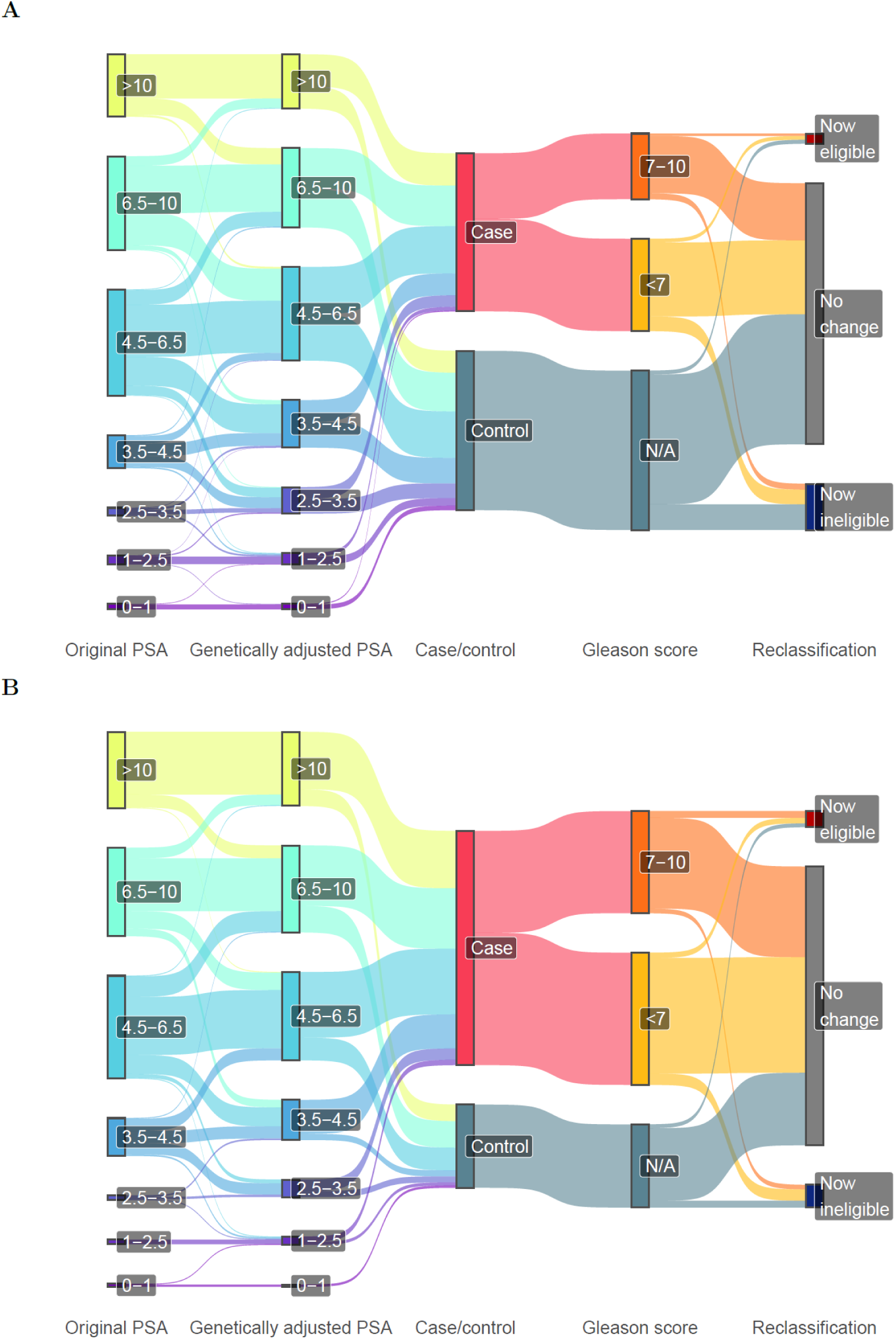
Biopsy reclassification with genetically-adjusted PSA. PSA levels were adjusted using the PRS-CSx estimate from the out-of-sample discovery cohort, assessed in GERA using age-specific cutoffs in **(A)** Europeans and **(B)** African Americans (see **Methods**). GERA Latinos and East Asians are shown in **Figure S3**.

To assess potential variability in genetic adjustment across PSA levels, we compared measured versus genetically adjusted PSA levels across a range of values in GERA men. Plotting measured versus adjusted PSA indicated consistent relative adjustment on the log scale (**Figure S5**). For example, at a measured PSA of 2.5, 6.5, and 10.0 ng/ml, the genetically adjusted PSA’ IQR ranged from 2.6-3.7, 4.6-6.8, and 9.1-12.5, respectively. These results suggest genetic adjustment is applicable to PSA values <20 displayed in the figure, although the implications are most profound around values where clinical decisions are made (e.g., near age-specific PSA thresholds).

### Impact of genetically-adjusted PSA on overall and aggressive PCa

Previous work has suggested that midlife PSA predicts lethal PCa^26^. In GERA European ancestry, genetically-adjusted midlife log PSA had a larger estimated magnitude of association with overall PCa (OR=4.57, 95% CI=4.27-4.88) than measured PSA (OR=4.30, 95% CI=4.04- 4.58)), though the CIs overlapped. The difference between associations was even larger for aggressive disease, with OR=3.92 (95% CI=3.54-4.35) for adjusted vs. OR=3.46 (95% CI=3.15- 3.81) for measured, though again CIs overlapped. African ancestry showed similar trends, with the genetically-adjusted association with PCa OR=5.85 (95% CI=4.73-7.23) vs. measured OR=4.72 (95% CI=3.56-6.27), and the aggressive genetically-adjusted OR=5.39 (95% CI=3.95- 7.35) vs. measured OR=4.72 (3.56-6.27). Estimates in Hispanic/Latino were also similar, but East Asian ancestry showed no difference (**Table S13**). Cross-validated AUC estimates also showed essentially no difference between adjusted and measured PSA, with estimates ranging from 0.7-0.8 in the different groups (**Table S13**).

### Associations with previously-reported PCa variants

In our discovery cohort, 20 of our 184 novel PSA-associated variants (10.8%) were genome-wide significantly associated with PCa in the PRACTICAL consortium’s European ancestry GWAS^27^ (**Tables S4, S6**). An additional 19 variants (10.3%) were associated with PCa at a Bonferroni level (p<0.05/184=0.00027). With correction for bias related to more frequent screening in men with higher constitutive PSA levels (see Methods)^20,28^, this count was reduced to 13 (7.0%) significant at the genome-wide level, and an additional 14 (7.6%) at the Bonferroni-corrected level. Out of the 111 novel PSA-associated variants from the meta-analysis, 8 (7.1%) were genome-wide significantly associated with PCa, and an additional 11 (9.8%) were significant at a Bonferroni level (p<0.00045). With bias correction, 5 (4.5%) were genome-wide significant, and an additional 4 (3.6%) were Bonferroni-significant.

### Associations with previously reported BPH variants

In our discovery cohort, one variant (rs1379553) was genome-wide significantly associated with BPH out of 137 variants available in a GWAS in UKB European ancestry^29^. An additional eight met a Bonferroni level (p<0.05/137=0.00036). Out of the 96 available variants identified from the meta-analysis, one variant was genome-wide significant (rs627320), and 6 more met a Bonferroni level (p<0.045/96=0.00052).

### Associations with urinary symptom variants

In variants identified in our discovery cohort, rs12573077 (p=8.4e-5) met a Bonferroni level (p<0.05/177=0.00028) for a test for an association with urinary symptoms in the GERA cohort (**Table S4, S6, S14**). In variants identified in the joint meta-analysis, none met a Bonferroni level of significance (p<0.05/110=0.00045).

### PSA variant associations with prostate volume

We found that 31 of the 407 PSA variants tested demonstrated some evidence of an association with prostate volume in the PASS cohort. rs182464120 on chromosome 19 was strongly associated (p=2.0e-11), rs12344353 on chromosome 9 was associated at a Bonferroni level (p=5.3e-5 <0.05/407=0.00012), and 29 other variants were associated at a nominal significance level (p<0.05) (**Table S15**).

### Associations with KLK3 plasma pQTL

We annotated the 447 variants from the joint GWAS meta-analysis using recently published plasma pQTL association results for *KLK3* from the UKB Pharma Proteomics Project using the Olink Explore platform;^30^ 409 had corresponding pQTL associations in the UKB. In European ancestry individuals (n=46,214), GWAS and *KLK3* pQTL effects were highly correlated (R=0.85, p=3.7e-117) (**Figure S6**). Eleven variants were associated with relative *KLK3* abundance at p<0.05/409, and, as expected, the strongest two associations were in *KLK3* (rs17632542, rs61752561) (**Table S16**). Among African ancestry individuals (n=1,065), we observed an attenuated correlation with effects on *KLK3* abundance (R=0.14, p=0.0034), although none of the individual pQTL associations reached statistical significance.

### Associations with eGenes

In our sc-RNA-seq data analysis, we found that each of the eGenes for PSA-associated variants is expressed across prostate cells. This is especially true in prostate luminal epithelial cells (which produce PSA) and is expected if the genes modify PSA levels (**Table S17**). **Figure S7** shows expression of the eQTL genes across multiple prostate tissue cell types, including luminal cells of the prostate epithelium and its precursor cells (e.g., basal epithelial cells of prostate; expression sorted by *KLK3*). Percentile expression of the eQTL-associated genes was significantly higher in luminal cells than all other prostate cell types (P=0.0006), suggesting these genes are more active in this cell type than other prostate cells (**Figure S8**) and supporting the hypothesis that these eQTL genes are involved in PSA expression.

## Discussion

Our GWAS detected 448 genome-wide significant variants associated with PSA levels, of which 295 were novel (184 in discovery and 111 in a meta-analysis), nearly quadrupling the total number of associated variants. The variance explained by genome-wide PRSs ranged from 11.6%-16.9% in men of European ancestry, 5.5%-9.5% in men of African ancestry, 13.5%- 18.6% in Hispanics/Latinos, and 8.6%-15.3% in the East Asian ancestry group. We also observed a decline in PRS predictive performance with increasing age, particularly at the oldest ages. The majority of newly identified variants were uniquely associated with PSA and not PCa.

Our discovery cohort included more African ancestry individuals than any prior study of PSA genetics. Of the eight genome-wide significant variants that were identified in the discovery phase in African ancestry, only two were sufficiently common to be assessed in European ancestry. Of those two, the association between rs1203888 (*LINC00261*) and PSA levels was unique to the African ancestry population. These eight variants generally failed to meet Bonferroni significance in our replication cohort, although the sample size was small (3,509 individuals of African ancestry); the variant rs18447639 in the *AR* gene was closest to replicating. Androgen receptor (AR) signaling is required for normal prostate development and function but is hijacked during carcinogenesis.^31^ Because prostate tumor growth and progression depend on AR signaling, androgen deprivation therapy remains a frontline treatment for progressing PCa, and the inhibition of *AR* activity may delay progression.^32^

A total of 10.8% of novel discovery and replication variants were prostate tissue eQTLs, and another 49.7% were eQTLs in other tissues. In addition, 16 discovery variants and five meta-analysis variants had predicted deleterious regulatory effects. Putative deleterious genes included: *AOC1*, which regulates histamine metabolism and sensitivity to non-steroidal anti- inflammatory drugs^33,34^; *MPZL2*, which is involved in thymus development and T-cell maturation; and *ZC3HC1*, a regulator of cell cycle progression and established susceptibility locus for coronary artery disease^35,36^. We also observed an association with PSA levels for the deltaF508 mutation in *CFTR* that causes cystic fibrosis, which is accompanied by infertility in 97% of affected males^37^ and has been linked to obstructive azoospermia (ClinVar^38^ accession SCV001860325). We detected another signal with possible links to male fertility, rs372203682 in *LMTK2*, a gene implicated in spermatogenesis^39^ that interacts with AR and inhibits its transcriptional activity^40^.

In SELECT, the variance in PSA levels explained by our independently associated GWAS variants was ∼1% larger than previously explained^20^ in European and ∼3% higher in African ancestry individuals. The variance explained in SELECT and PCPT was substantially less than that in GERA, even though we evaluated only variants from our discovery cohort (which did not include GERA). This may be due in part to the studies’ selection criteria, as individuals in SELECT and PCPT were required to have PSA≤3 ng/mL^23^ and ≤4 ng/mL^24^, respectively, at baseline. However, in AOU, which did not have this selection criteria, variance explained for European ancestry men was at most 0.5% higher than SELECT, and thus also lower than GERA. For African ancestry men in AOU, variance explained was 2-3% lower than in SELECT, suggesting that differences in performance may be attributed to factors other than preferential selection for low baseline PSA. We also investigated whether BPH may contribute to variability in PRSs performance. The estimated variance explained was <0.5% higher when excluding men with a BPH diagnosis. By BPH, we mean a clinical diagnosis rather than presence of BPH. Most patients evaluated for potential PCa have evidence of BPH, which can result in elevated PSA levels^41,42^. These findings highlight the need to evaluate genetically adjusted PSA in a wider range of clinical settings, as well as the challenges with curating out-of-sample cohorts with clinical data sufficient for such evaluations.

The performance of PRS constructed using weights derived from the multi-ancestry meta-analysis typically matched or surpassed the performance of PRSs based on ancestry-specific weights. As expected, genome-wide PRS-CSx generally achieved 1-6% higher explained variance than the PRSs limited to mJAM genome-wide significant variants. However, the improvement in performance observed for PRS-CSx was not equal across populations. The largest increase was observed for Hispanic/Latino men, followed by European ancestry men (generally similar to Hispanic/Latino men), followed by Asian ancestry men. To our knowledge, this is the first time out of sample PRS performance was assessed in a Hispanic/Latino population. Relative to the fine-mapped PRS, the degree of improvement was smallest for African ancestry. This may be due to a number of factors. First, PRS-CSx uses a single hyperparameter to couple posterior effect sizes across ancestry groups, which may be insufficient to capture different correlation structures among populations. Second, HapMap3 variants used by PRS-CSx do not tag genetic variation equally well across ancestries. Fine-mapping PRS methods do not limit to this set of tagging SNPs and may be more likely to capture population-specific variants. Third, the choice of linkage disequilibrium (LD) reference panels has slightly different implications for the two PRS approaches. PRS-CSx relies on LD reference panels for estimating joint SNP effect sizes, while fine-mapping requires LD information for identifying independent variants from summary statistics. mJAM advances other fine-mapping approaches by incorporating population-specific LD, which is more accurate than using a single population as the LD reference^21^ or only making use of the largest ancestry group. While PRS-CSx provides more flexibility to accommodate different genetic architectures, it may be more sensitive to the choice of LD reference panels and mismatches in LD structure between PRS training and testing populations, especially without a separate dataset for parameter tuning.

We found that genetically adjusting PSA levels reduced unnecessary biopsies in controls, albeit less so than in previous work^20^ in the same GERA population. It is likely that our previous study overestimated reclassification in controls because there was partial overlap between the GWAS meta-analysis used to train the PRS used for adjustment - GERA was included - and the population in which we undertook genetic adjustment. In the present study, we performed genetic adjustment using a PRS trained on a large GWAS that did not include GERA. We also saw an increase in magnitude in genetically-adjusted midlife PSA association with PCa in most GERA groups, although CIs overlapped for all, and while our previous study^20^ did not see any benefit in African ancestry, here we saw an increase.

Our investigation had several limitations. Relative to prior studies of PSA genetics, the discovery and replication cohorts included here substantially increased the number of men from diverse populations. While both were very large (N∼300K and ∼100K men total), the replication cohorts had disproportionately smaller African ancestry (discovery N ∼58K and replication N ∼3.5K) and Hispanic/Latino populations (N∼24K and ∼3K). Going forward, our PSA Consortium will continue to seek new study populations with both genotypic and phenotypic data that represent diverse participants. Nevertheless, for African ancestry, 43% of variants met a nominal replication threshold, many more than the 5% that would be expected by chance. We also suspect that we had limited power to detect effect size heterogeneity, especially since variants that exhibited significant heterogeneity were mostly known variants in strongly associated regions. Another limitation is that GERA biopsy reclassification may have been specific to Kaiser Permanente clinical guidelines, as previously discussed^20^. In addition, while we did our best to restrict relevant analyses to PCa-free individuals, some individuals likely had undetected PCa^43^.

However, the number was unlikely to be large enough to materially impact our results because our study population was relatively young; the average age among men in the MVP (which comprised a large majority of our discovery study population, N∼286K) was 58 years for men of European ancestry, 52 years for men of African ancestry, and 54 years for Hispanic/Latino men or Asian ancestry men. Further, the PSA PRS_CSx_ explained increasing variability in PSA levels for individuals of younger ages. Most novel PSA-associated variants were not associated with PCa, and those that were may have been due to screening bias, as previously shown^20^. The lack of BPH information in most of our cohorts was an additional limitation, but most novel variants associated with PSA levels were not associated with BPH in others’ work on UKB European ancestry individuals^29^, and the variance explained by PRSs in SELECT was affected by <0.5% in participants with BPH. We were unable to account for prostate volume, a strong predictor of PSA levels^44^. Finally, we note that our GWAS and resulting PRSs were developed for total PSA. Future work should work toward capturing genetic factors that are specific to constituents of total PSA.

In summary, we undertook a large-scale, multi-ancestry study with over three times the sample size of previous work,^20^ expanded our understanding of the genetic basis of PSA levels and our potential to improve the accuracy of PSA genetic adjustment across ancestries. Using an ancestrally diverse study population, we detected hundreds of novel variants associated with PSA levels that were largely independent of PCa and BPH. These findings explain additional variation in PSA levels, especially among men of African ancestry, who suffer the highest morbidity and mortality due to PCa, as well as among Hispanic/Latino men. This highlights the importance of studying diverse populations to enable novel discoveries and construct PRS that will perform equally across ancestry groups. Taken together, our work moved us closer to leveraging genetic information to personalize and substantially improved our understanding of the genetic basis of PSA levels and of genetic adjustment of PSA levels across individuals of diverse ancestries.

## Methods

### Discovery participants and phenotype measurements

Our primary analyses included 296,754 men from seven cohorts that had not previously been analyzed in studies of PSA genetics. These are described briefly below; additional details, including array, ancestry, imputation reference panels, sample sizes, number of variants, and standard filters applied are described in **Tables S1-S3**. To ensure participants had a functional prostate unaffected by surgery or radiation and to exclude individuals at a high risk of undiagnosed PCa^45^, participants were restricted to men with no history of PCa or surgical resections of the prostate, and at least one PSA measurement between 0.01 and 10ng/mL. Analyses were based on each individual’s earliest recorded PSA level. For descriptive statistics, meta-analysis of PSA medians from each cohort was done with the weighted median of medians method in the R v4.2.3^46^ package metamediation v1.0.0^47^. Sub-populations were defined by self-identified race/ethnicity and/or genetically-inferred ancestry, depending on the cohort.

*African American Prostate Consortium (AAPC)*. The AAPC is comprised of African ancestry studies with PCa phenotyping.^27^

*Mount Sinai BioMe® Biobank (BioMe)*. BioMe is a longitudinal cohort linked to Epic electronic health records (EHRs)^48^. Individuals were European ancestry, Hispanic/Latino, and African ancestry.

*Chicago Multiethnic Prevention and Surveillance Study (COMPASS)*. COMPASS is a longitudinal study of Chicagoans with >11,000 participants currently enrolled (82% African American).^49^ PSA data has been described previously^50^.

*Men of African Descent and Carcinoma of the Prostate (MADCaP)*. MADCaP is a consortium of epidemiologic studies addressing the high PCa burden in African ancestry men.^51,52^

*Multiethnic Cohort (MEC)*. MEC is a prospective cohort study that enrolled >215,000 Hawaii/Los Angeles residents ages 45-75 years between 1993-1996.^53,54^

*Million Veteran Program (MVP)*. MVP is a multi-ancestry cohort recruited nationwide. Information is obtained from EHRs, including inpatient International Classification of Diseases (ICD)-9 codes, Current Procedural Terminology (CPT) procedure codes, clinical laboratory measurements, and reports of diagnostic imaging modalities^55^. Sub-populations were created using the harmonized ancestry and race/ethnicity (HARE) method.^56^

*Southern Community Cohort Study (SCCS)*. SCCS is a prospective cohort study that recruited 85,000 predominantly African ancestry adults from community health centers in the southeastern United States. This study included only men of African ancestry.^57^

### Replication cohorts

Genome-wide significant variants identified in the discovery cohort were tested for replication in the previous largest GWAS of PSA levels, which included 95,768 men (85,824 European ancestry, 89.6%)^20^, using a Bonferroni corrected α level. In addition, previously-identified genome-wide significant variants ^20^ were tested for replication in our independent discovery cohort. Statistical tests throughout were two-sided.

### Additional PRS evaluation cohorts

For our discovery cohort results, we evaluated PSA PRS performance and reclassification in individuals from the GERA cohort (also in the replication cohort, out-of-sample for the discovery cohort (n=35,322; 28,503 European; 2,716 Latino; 2,518 East Asian; and 1,585 African American)).

Additional out-of-sample cohorts for (both the discovery analysis and the joint meta-analysis of discovery and replication) PRS assessment was done in genotyped individuals from the PCPT^24^ (n=5,725 European) and SELECT^23^ (n=25,366; 22,173 European; 1,763 African American/European; 1,173 African American; and 257 East Asian) and All of Us (AOU; n=17,512; 11,922 European; 2,469 African American; 1,783 other; 1,336 Hispanic/Latino)^25^, which have been previously described. Briefly, PCPT and SELECT began as randomized, placebo-controlled, double-blind clinical trials of finasteride and selenium and vitamin E, respectively, and both enrolled men ≥55y. Individuals in SELECT and PCPT were required to have PSA≤3 ng/mL^23^ and ≤4 ng/mL^24^, respectively, at baseline. The National Institutes of Health (NIH) AOU cohort is committed to including groups that have been historically underrepresented in research^25^. From AOU, we selected individuals with PSA>0.01 between the ages of 40 and 90, with short-read whole-genome sequencing (WGS) data and no survey or EHR conditions/observations reflecting a history of PCa. The median PSA measurement we used was required to be ≤10 ng/mL. PRSs were calculated with the WGS data subset to variants with population-specific allele frequency ≥1% or a population-specific allele count >100 for any genetic ancestry. Genetic ancestry was determined using a random forest classifier trained on the principal component (PC) space of the Human Genome Diversity Project and 1000 Genomes Project (KGP)^58^.

### Ethical considerations

Informed consent was obtained from all study participants. AAPC was approved by their IRB. The ethics review board of the Program for the Protection of Human Subjects of Mount Sinai School of Medicine approved BioMe (#HSD09-00030, #07-0529 0001 02 ME). The University of Chicago Biological Sciences Division IRB Committee A (#IRB12-1660) approved COMPASS. Local and national IRBs approved MadCAP. MEC was approved by their IRB. The VA Central institutional review board (IRB) approved the MVP. The IRBs at Vanderbilt University and Meharry Medical College approved SCCS. Vanderbilt University Medical Center IRB approved BioVU. GERA was approved by the Kaiser Permanente Northern California IRB and the University of California, San Francisco. A local ethics committee approved the Malmo Diet and Cancer Study (MDCS). The Prostate, Lung, Colorectal and Ovarian (PLCO) Cancer Screening Trial was approved by the IRBs at each participating center and the National Cancer Institute, and the informed consent document allows data use for cancer and other adult disease investigations; we used publicly posted summary statistics, for which no IRB is required. The research was conducted with approved access to UKB data (#14105).

### Genotype quality control and imputation

Study subjects were genotyped using conventional GWAS arrays (**Table S1**). Genotypes were then imputed using imputation servers^59^, Minimac3^60^, or IMPUTE2^61^. The vast majority of studies imputed to the KGP phase 3 reference panel^62^, with one sub-study imputing to KGP phase 1 just for the X chromosome^63^ and another imputing to the TOPMed reference panel^59^. Since all but two studies (>95% of participants) used genome build 37, welifted over the assembly of those from build 38 using triple-liftOver^64^ v133 (2022-05-20), an extension of LiftOver^65^ that accounts for regions that are inverted between builds.

Standard genotype and individual-level QC procedures were implemented in each ancestry group in each participating study. Specific study protocols are delineated in **Table S1**, with additional QC steps and details in **Table S2**. Unless information was unavailable or a filter did not make sense for a particular group, variants were retained if their imputation quality score was ≥0.3, their MAF was ≥0.5% if the sample size was ≥1000 and ≥5% otherwise, their Hardy-Weinberg equilibrium (HWE) was ≥1e-8, they were mapped in build 37, and they had an MAF difference ≤0.2 compared to KGP populations (full details in **Table S3**). For the cohorts that meta-analyzed sub-cohorts (e.g., the three small African ancestry sub-cohorts within the SCCS African ancestry group; **Table S2**), we also required that variants be present in all sub-cohorts (necessary for multi-ancestry analysis method limitations, although this removed only a very small number of variants, **Table S3**). Finally, we excluded variants if they were present in only one study with n<2,000.

### Association analyses

GWAS within each ancestry group in each study were undertaken using linear regression of log PSA on additive genotypes, and, when using multiple measurements, the long-term average residual by individual^66^. The minimum set of covariates included age at PSA measurement and genetic ancestry PCs. If available, GWAS also adjusted for batch/array, body mass index, and smoking status (**Table S1**). Meta-analyses of each ancestry group and across the overall discovery cohort were conducted using inverse-variance weighted fixed effects models using a custom patched version of METAL v2011-03-25 that prevents numerical precision loss (lines 633 and 635 of “Main.cpp” modified to the number 15 to output 15 digits precision)^67^. We also assessed heterogeneity with Cochran’s Q across the four ancestry groups.

To identify independently associated genome-wide significant (p≤5e-8) variants with computational efficiency, we first formed clumps of genome-wide significant variants such that all clumps were ≥10Mb apart and independent of one another; specifically, the top variant was chosen, genome-wide significant variants ≤10Mb from any variant in the clump were added to the clump, the process was iterated until a final clump was formed, and then the process was repeated to form more clumps (i.e., clumps were created such that there was no additional genome-wide significant variant ≤10Mb). Within each clump, we used mJAM v2022-08-05^21^, which uses population-specific LD reference panels for each contributing cohort and ancestry group to model the correlation among variants, with an r^2^<0.01 threshold in all ancestry groups. Genotypes utilizing the appropriate GERA cohort group (European, Hispanic/Latino, African American, and East Asian) served as references^68^.

To maximize discovery efforts, we combined our discovery cohort (n=296,754) with our replication cohort (n=95,768), for a total of 392,522 individuals.

Associations were considered novel if they had low LD from all previously-reported variants^20^. Specifically, we required r^2^<0.01 in all four ancestry groups, again using GERA as LD reference.

### Annotation

Variants were annotated using FUMA^69^. We first prioritized genes that included a significant prostate eQTL from GTeX v8 (www.gtexportal.org). We then prioritized other significant eQTLs and finally by closest gene. Deleteriousness of mutations was determined by CADD scores; a recommended cutoff to identify potentially pathogenic variants of scores ≥15 has been suggested (the median of splice site changes and non-synonymous variants from CADD v1.0; corresponds to the top 3.2% of variants)^70^. Gene functions were characterized with RefSeq^71^. Circos plots were generated using Circos v0.69-6^72^.

### Medication sensitivity analysis

Some of our study participants may have taken medications that can affect PSA levels. In particular, 5-alpha reductase inhibitors and testosterone can impact PSA levels^73,74^. We assessed the use of these medications among 26,669 men of European ancestry in the UKB with at least one PSA measurement. Men with a prescription for at least one of the two medications prior to PSA measurement were considered users. Ten percent of the men were prescribed 5-alpha reductase inhibitors and 0.56% testosterone. We also controlled for potential confounding by alpha blocker use.

### Out-of-sample PRS variance explained

We calculated PRSs to assess the overall PSA variance explained by genetics, and to adjust PSA measurements for PSA genetics. All PRS results are shown only in independent cohorts (i.e., training dataset completely independent of testing dataset), such that assessments of performance are unbiased. Nonparametric bootstrap percentile CIs for variance explained were calculated using 1000 replicates.

We used two sets of individuals to construct the PRSs. First, we constructed PRSs from our discovery cohorts to allow assessment in GERA, PCPT, SELECT, and AoU. Second, we constructed PRSs from the meta-analysis of discovery and replication cohorts (which included GERA), with assessment in PCPT, SELECT, and AoU only. For GERA we included results using first and multiple measurements; for PCPT and SELECT we include results using the first measurement.

We also used two sets of variants to calculate the PRSs in each of the two sets of individuals. We first utilized the independent genome-wide significant variants discovered in our analyses (one for discovery and one for the meta-analysis of discovery and replication). Second, we constructed a genome-wide score using PRS-CSx v2023-08-10^75^, which was implemented utilizing GWAS summary statistics, the 1,287,078 HapMap3 variants as an LD reference that had an imputation quality >0.9 in SELECT, using a global shrinkage parameter of ϕ=0.0001 (which performed well in our previous work^20^), and variants with imputation quality ≥0.9. Since PRS-CSx only considers autosomes, independent genome-wide significant X chromosome variants were included (and produced a negligible increase in performance). The final scores were calculated by summing the effect size times the (probabilistic) number of alleles at each locus with PLINK v2.00a3.7LM^76^.

We also assessed the variance explained by the discovery PRS-CSx within age intervals in GERA; we looked only in GERA to have an out-of-sample estimate from discovery and a large enough sample size at each age. An individual could be in multiple bins, but using just the first measurement of that individual per age bin.

### Fur PSA for PCa screening in GERA

We adjusted PSA levels as has been described previously^20^. Briefly, PSA values for individual i were adjusted by PSA_i_^adj^ = PSA_i_ / a_i_, where a_i_ is a personalized adjustment factor derived from our PRS, as: a_i_ = exp(PRS_i_) / exp(mean(PRS)). Here we estimated the mean(PRS) value within each group in the GERA cohort. We then evaluated the potential utility to alter biopsy referrals using age-specific PSA thresholds used within the Kaiser system (40-49y=2.5, 50-59y=3.5, 60- 69=4.5, and 70-79=6.5 ng/ml^77^), evaluating net reclassification in cases and controls^20^.

We also tested for associations of our PSA^adj^ with Gleason score (≤6, 7, and ≥8) using multinomial logistic regression with the R package nnet v7.3.18^78^.

To assess whether there was variability in PSA adjustment across PSA levels, we first binned PSA values (with smaller ranges in lower values where there was more data). Within each bin, we computed PSA - PSA_adjusted_, and then computed the median and IQR of these values. The median and IQR were then plotted at the center point of each bin by adding them to the identity line.

#### Impact of genetically-adjusted midlife PSA on overall and aggressive PCa prediction

We next investigated the impact of genetically adjusting PSA on the prediction of overall and aggressive PCa in the GERA cohort (3540 cases [1,028 aggressive, Gleason ≥7], 21,702 controls). We constructed a midlife PSA^26^ based on each participant’s median PSA between 50- 60y, with cases restricted to measurements ≥1y before diagnosis. Genetic PSA adjustment was performed as in the previous section. Associations between PSA or genetically adjusted log PSA and PCa risk were assessed using logistic regression for overall PCa cases vs controls and for aggressive cases vs. controls, adjusting for covariates in Table S1. AUC was estimated using 10-fold repeated cross-validation (10 repeats) with caret v6.0.90^79^.

### Bias-corrected PCa estimates

PCa associations in individuals with European ancestry in the PRACTICAL consortium^27^ were adjusted for screening bias^28^, using estimates previously derived^20^: β’_Cancer_ = β_Cancer_ - bβ_PSA_, SE’_Cancer_=(SE_Cancer_^2^ + b^2^SE_PSA_^2^ + SE ^2^β_PSA_ + SE ^2^SE_PSA_^2^), where SE is the standard error, and estimates were b=1.144, and SE_b_=2.909e-4.

### Associations with urinary symptom variants

We evaluated whether the novel PSA variants were associated with urinary symptoms in the GERA cohort. Individuals in this cohort completed the first 6 (of 7) questions from the American Urological Association Symptom Index (AUA-SI)^80^ with 5-point Likert scale responses. The questions asked about incomplete emptying, frequency, intermittency, urgency, weak stream, and straining (**Table S13**). The one missing question from the AUA-SI regarded nocturia. We calculated total scores as the sum of the questions, giving each individual a value ranging from 6-30. The score was dichotomized at <7, ≥7 to differentiate men with little or no BPH (N=12,846) from those with moderate or severe BPH (N=15,480). We then assessed the association between the PSA variants and the urinary symptom score.

### Prostate volume analysis

We evaluated associations between PSA variants and prostate volume in patients on active surveillance (AS) enrolled in the Canary Prostate Active Surveillance Study (PASS). Between 2008-2017, PASS prospectively enrolled 1455 patients with clinically localized PCa (cT1-cT2 and Gleason Grade 1-2) to undergo AS at one of 10 national sites^81^. Prostate volume was measured at diagnosis, with a median measurement of 43.0cc (IQR=31.0-57.5). The median age at diagnosis was 63 years (IQR=58-67), and 85% of the PASS cohort self-reported as European ancestry. Genotyping was conducted in 1,220 participants^82^. We assessed potential associations between the 407 PSA variants that we successfully imputed in PASS and prostate volume using mixed models with fixed effects for genetic variants, age at diagnosis, and 10 PCs, and a random effect for a genetic relationship matrix.

### Associations with eGenes

We used single cell RNA sequencing (scRNA-seq) data to assess whether the eGenes for PSA-associated variants (Table S7) are expressed in secretory prostate cell types, particularly luminal epithelial cells, more so than other prostate cell types. For these analyses, we used data from the Chan Zuckerberg Cell by Gene census v2023-12-15^83^.

## Data availability

Summary statistics (from the discovery analysis and the final meta-analysis) will be made available in the NHGRI-EBI GWAS Catalog (https://www.ebi.ac.uk/gwas/downloads/summary-statistics). PRS weights (PRS_318_, PRS_447_, PRS_CSx,disc_, PRS_CSx,joint_) will be made available in the PGS catalog (https://www.pgscatalog.org/). To protect individuals’ privacy, complete GERA data are available upon approved applications to the Kaiser Permanente Research Bank Portal (https://researchbank.kaiserpermanente.org/for-researchers). UKB data are available in the UKB cloud-based Research Analysis Platform (https://www.ukbiobank.ac.uk). GTEx data were obtained from the GTEx portal (www.gtexportal.org) and can be obtained from dbGaP Accession phs000424.v8.p2.

## Code availability

Genome-wide association analysis were conducted using PLINK v2.0a3.7LM (http://www.cog-genomics.org/plink/2.0/). Meta-analysis were conducted with a custom-patched METAL v2011-03-25 (https://genome.sph.umich.edu/wiki/METAL_Documentation) that prevents numerical precision loss (lines 633 and 635 of “Main.cpp” modified to the number 15 to output 15 digits precision”), and with MJAM v2022-08-05 (https://github.com/USCbiostats/hJAM/R). Imputation was done via imputation servers (https://imputationserver.sph.umich.edu, https://imputation.biodatacatalyst.nhlbi.nih.gov), Minimac3 (https://genome.sph.umich.edu/wiki/Minimac3), and IMPUTE2 (https://mathgen.stats.ox.ac.uk/impute/impute_v2.html). Analysis were also conducted in R, including v4.2.0 (https://cran.r-project.org/). FUMA was used for annotation (https://fuma.ctglab.nl). Circos plots were generated using Circos v0.69-6 (https://fuma.ctglab.nl). The genome-wide PRS was conducted with PRS-CSx v2023-08-10 (https://github.com/getian107/PRScsx).

## Data Availability

Summary statistics (from the discovery analysis and the final meta-analysis) will be made available in the NHGRI-EBI GWAS Catalog (https://www.ebi.ac.uk/gwas/downloads/summary-statistics), and PRS weights (PRS318, PRS447, PRSCSx,disc, PRSCSx,joint) in the PGS catalog (https://www.pgscatalog.org/). To protect individuals privacy, complete GERA data are available upon approved applications to the Kaiser Permanente Research Bank Portal (https://researchbank.kaiserpermanente.org/for-researchers). UK Biobank data are publicly available by request from https://www.ukbiobank.ac.uk. GTEx data was obtained from the GTEx portal (www.gtexportal.org) and can be obtained from dbGaP Accession phs000424.v8.p2.

## Acknowledgements

We thank the participants who generously agreed to participate in each cohort. This research is based in part on data from the Million Veteran Program (MVP), Office of Research and Development, Veterans Health Administration, and was supported by the MVP017 Exemplar Cancer Project. This research was conducted using the UKB resource under application number 14105. In addition, the All of Us Research Program would not be possible without the partnership of its participants.

